# The effect of potential factors on all-cause and cause-specific and mortality: a pre-COVID-19 period review

**DOI:** 10.1101/2022.06.08.22276123

**Authors:** Jia Xu, Yunkai Lin, Jianyu Tong, Ying Zhou

**Author notes:** **Corresponding author:** Ying Zhou, PhD, School of Public Health, Shenzhen University, Shenzhen, China.

## Abstract

**Background:** About 3 million people die every year in USA. In order to provide a general direction and background for subsequent research and development in this field, we reviewed the studies about impact of potential factors associated with the main causes of mortality and all-cause mortality.

**Method:** Researches are selected by PubMed website for last 15 years with published language of English. The leading cause of death were published by CDC in 2020 (excluded COVID-19) including the ten natural mortalities and unnatural mortality. We summarized the potential factors associated with the mortality and sorted them by “positive/negative effect” and “long-term/short-term effect”.

**Result:** Among risk factors, the factors of depression, unhealthy diet, overweight and obesity and other similar factors increase the mortality of main leading natural causes. Among protective factors, the factors of physical activity, nut intake, chocolate consumption were associated with the reduced mortality of multiple diseases. We also found that some factors possess bidirectional influence on different diseases or even one disease. The gender of female negatively affects mortality of diabetes, but positively affects cardiovascular diseases. The majority of air pollutants are risk factors for respiratory diseases while ozone possibly reduce the mortality.

**Conclusion:** Our review summarized various factors which have positive/negative and long-term/short-term effects on the main mortality of cause-specific and all-caused mortality. Further study is required to investigate the contribution of different variable on mortality.

## Introduction

Many diseases, such as heart disease, respiratory infection and cancer, can result in great harm to human health. The patients not only have to suffer from the financial burden, but also have to endure the dramatically decline of life quality. Some diseases will even lead to death when the syndrome become grievous. According to the WHO investigation, In 2019, the top 10 leading causes of death deaths for 55% of the 55.4 million Deaths worldwide(1). Meanwhile, some kind of deaths, like heart disease, cancer, unintentional injuries, chronic lower respiratory disease, stroke, Alzheimer’s disease, diabetes, kidney disease, influenza and pneumonia, and suicide accounted for 73.4% of all deaths in the United States(2). Therefore, investigation of factors associated with all-cause and cause-specific mortality is of great importance.

Plenty of previous studies have evaluated the associations between different causes of mortality and their potential factors. Causes of mortality such as cardiovascular disease, chronic kidney disease and cancer, have been found much correlations between many risk factors(3-5). Some risk factors such as obesity, alcohol consumption and physical activity have also been demonstrated associations with various causes of mortality(6-8). On the other hand, specific populations, for instance, children and adolescents(9), the elderly(≥65 years)(10) have different relationships between all-cause mortality and their risk factors. Several studies investigated the potential factors and various mortality in different regions, for example, Kenya summarized risk factors of all-cause mortality at the national and county levels(11). However, most researches are limited to exploring the relationship between only mortality from one or a few specific causes and risk factors. Current related reviews either only focus on a few of risk factors, or include fewer cause-specific mortality, or focus on a specific region and population, which have some particular limitations. It is necessary to summarize more potential factors associated with all-cause and cause-specific mortality.

The aim of our study is to systematically review previous published researches, extract potential factors associated with all-cause and cause-specific mortality. We chose the main leading causes of deaths in the USA and searched the their potential factors in Pubmed dataset. By summarizing multiple potential factors, our paper aims to make a macro analysis and describe the impact of long-term or short-term factors, together with risk and protective factors in a comprehensive and detailed way, which expecting to provide a general direction and background for subsequent research and development in this field.

## Materials and methods

### 1. Search strategy

Our review is to research the relationship between some factors and cause-specific death in pre-COVID-19 period time so the factor of COVID-19 is excluded. A search was performed on PubMed websites to screen for relevant articles published in the previous 15 years (Table 1). We used the top ten causes of death published by CDC in 2020 and excluded death of COVID-19. We added the Septicemia and defined “unnatural cause” as the mortality of “all cause” subtracts “natural cause”. As the result, the specific causes we concerned contained Diabetes mellitus, Cerebrovascular diseases, Alzheimer’s diseases, Heart diseases, Nephritis, nephrotic syndrome, and nephrosis, Unnatural cause (accident and suicide), Septicemia, Neoplasm, Influenza and pneumonia, Other Respiratory diseases (Table 2). The selected articles included possible factors of cause-specific or all-cause mortality at the population levels.

**Table 1.**
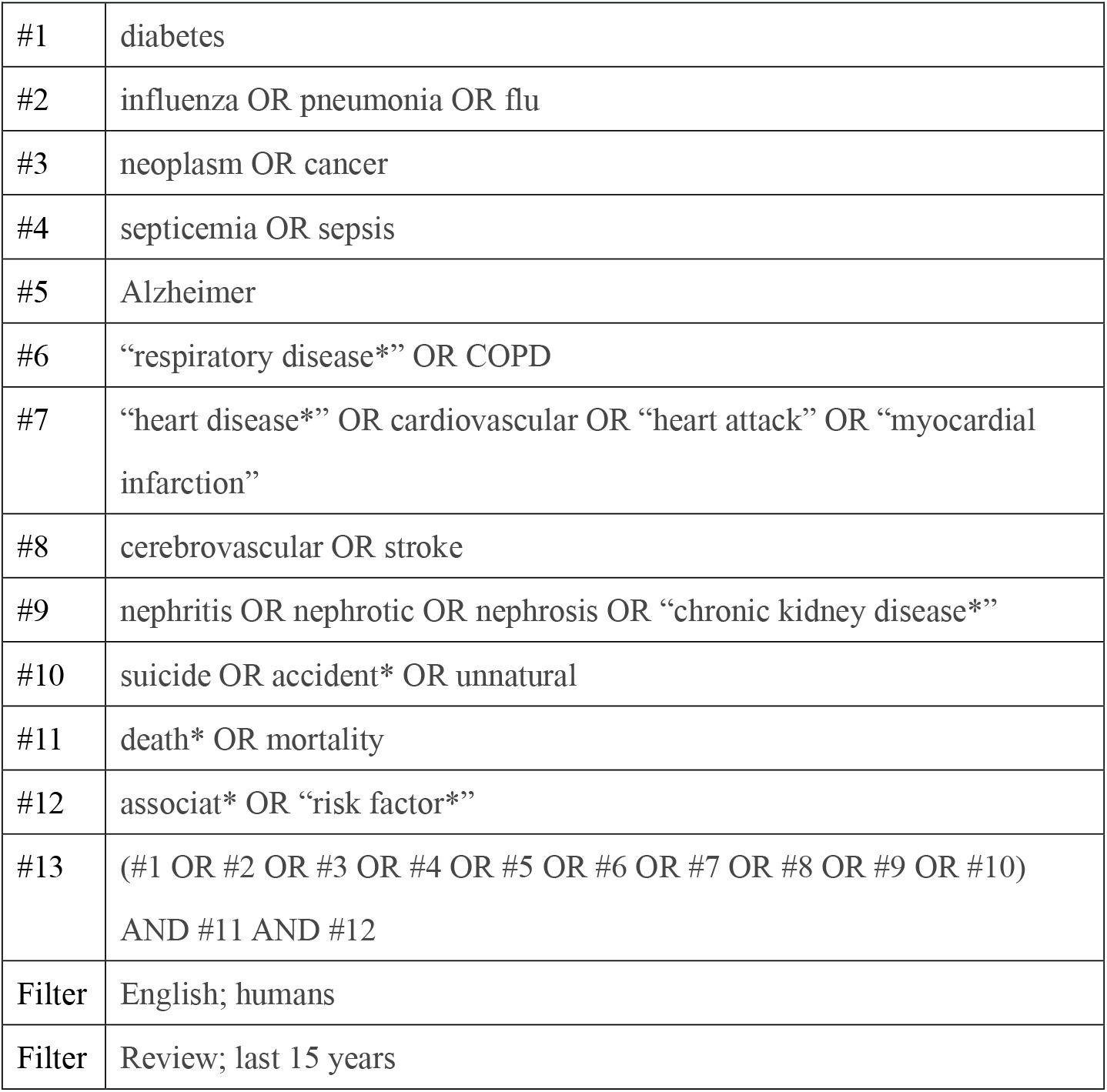
PubMed search terms and applied filters.

**Table 2.** ICD-10 code for cause-specific mortality.

| **Cause-specific Mortality** | **ICD-10 code** |
| --- | --- |
| Diabetes mellitus | E10-E14 |
| Cerebrovascular diseases | I60-I69 |
| Alzheimer's diseases | G30 |
| Heart diseases | I00-I09, I11, I13, I20-I51 |
| Nephritis, nephrotic syndrome, and nephrosis | N00-N07, N17-N19, N25-N27 |
| Unnatural cause (accident/suicide) | / |
| Septicemia | A40-A41 |
| Neoplasm | C00-C97 |
| Influenza and pneumonia | J09-J18 |
| Other Respiratory diseases (Chronic lower respiratory diseases and other diseases of respiratory system) | J00-J06, J30-J47, J67, J70-J98 |

### 2. Inclusion and exclusion criteria

A series of inclusion and exclusion criteria about the filter of paper heading, keywords and abstract to make sure the studies can be selected into our review. Due to the large amount of search results, filters were applied to select for reviews, English language articles, and articles on human species. The search retrieved 11,715 articles. The titles and abstracts of the reviews were screened for relevance independently by two independent researchers and additional articles cited in reviews were also considered. The search was performed with terms that included all specific causes in combination with death and mortality. All kinds of reviews can be included but not limited to narrative, scoping, and systematic reviews.

The exclusion criteria involved articles that were focused on clinical treatments or molecular mechanisms. All type of articles which were included how the factor of COVID-19 influence the specific diseases cannot be selected because our review is to research the pre-COVID-19 period. Besides, the qualitative research studies were also excluded.

### 3. Articles sorting

In consideration of a wide range of potential factors, there are 21 specific factors included anxiety, depression, air pollution, smoking, drinking alcohol, obesity, social support, nut intake, coffee consumption, chocolate consumption, green tea consumption, nutrition, physical activity, medical resource(including medical attention delayed, social economic status about medical care, screening, diagnosis, and treatment), high blood pressure, sunlight, napping, sleep, diet, ICU (including ICU care, admission to ICU, Emergency department to ICU Time), gender, kidney function. We sorted related articles to clearly show the positive or negative and long-time or short-time factors of the mortality from specific diseases. A list of other factors are provided in Additional file: Table S1. As the various factors can affect specific diseases within different time period, we sorted factors into two groups according to the time lag between the factors and mortality. We defined”short-term factors’’ as time lag of less than one month and”long-term factors” on the contrary.

## Results

We have searched total 11715 studies from PubMed website according our methods and finally obtained 97 articles which met our inclusion and exclusion criteria. The main potential factors which could positively or negatively affected the all-cause and cause-specific mortality was displayed in the figure 2 (Protective and risk factors). We also divided these main factors into two types of long-term and short-term factors in Figure 3.

**Figure 1.**
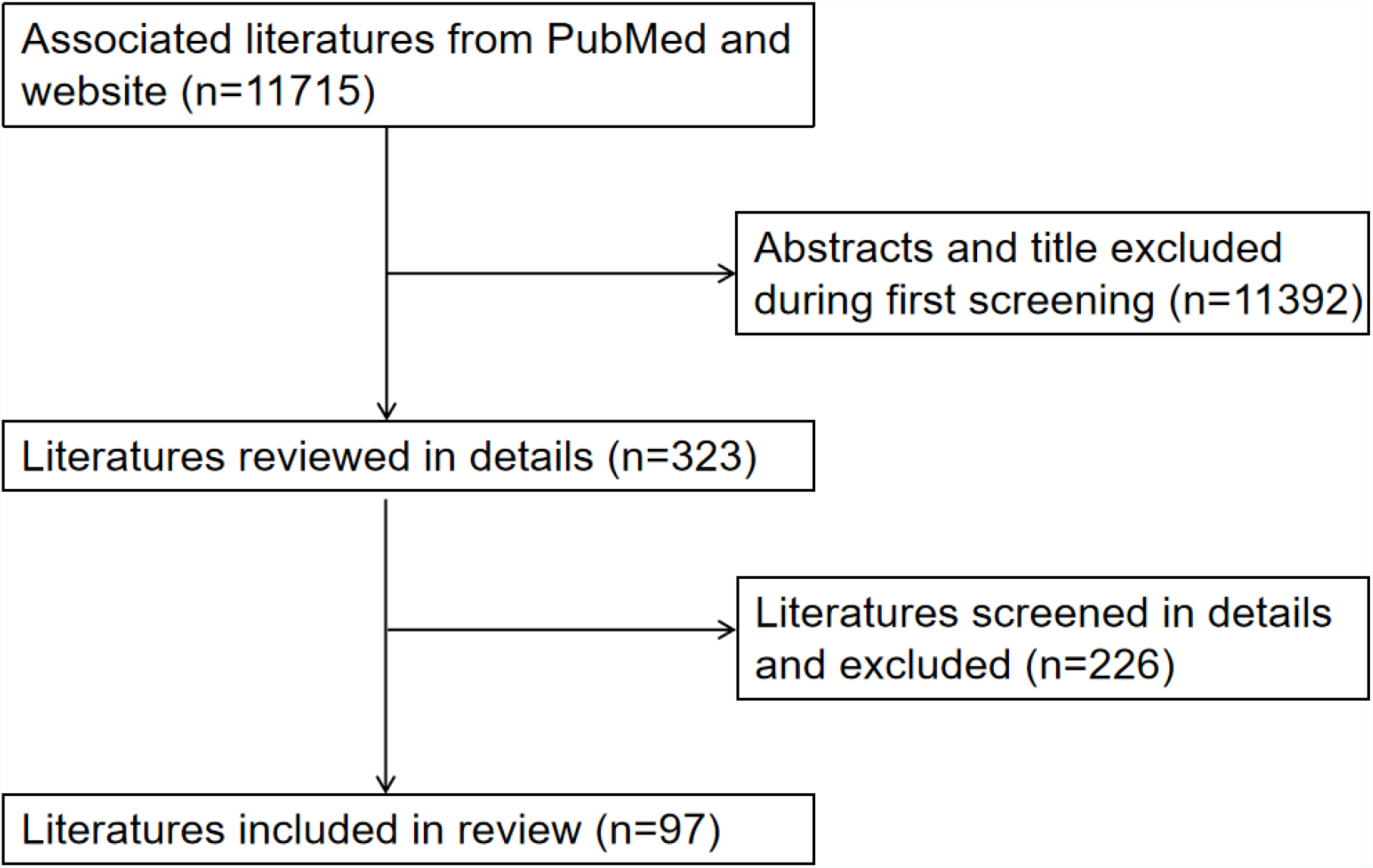
Flow diagram of the literature search and researches filter process.

**Figure 2.**
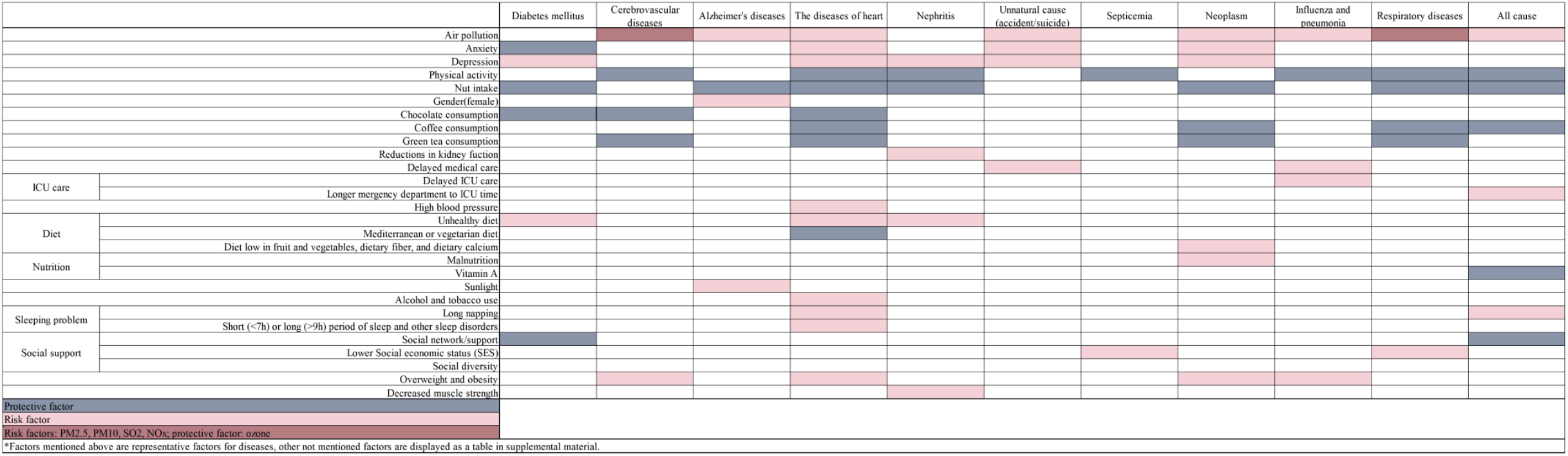
Protective and risk factors associated with cause-specific mortality.

**Figure 3.**
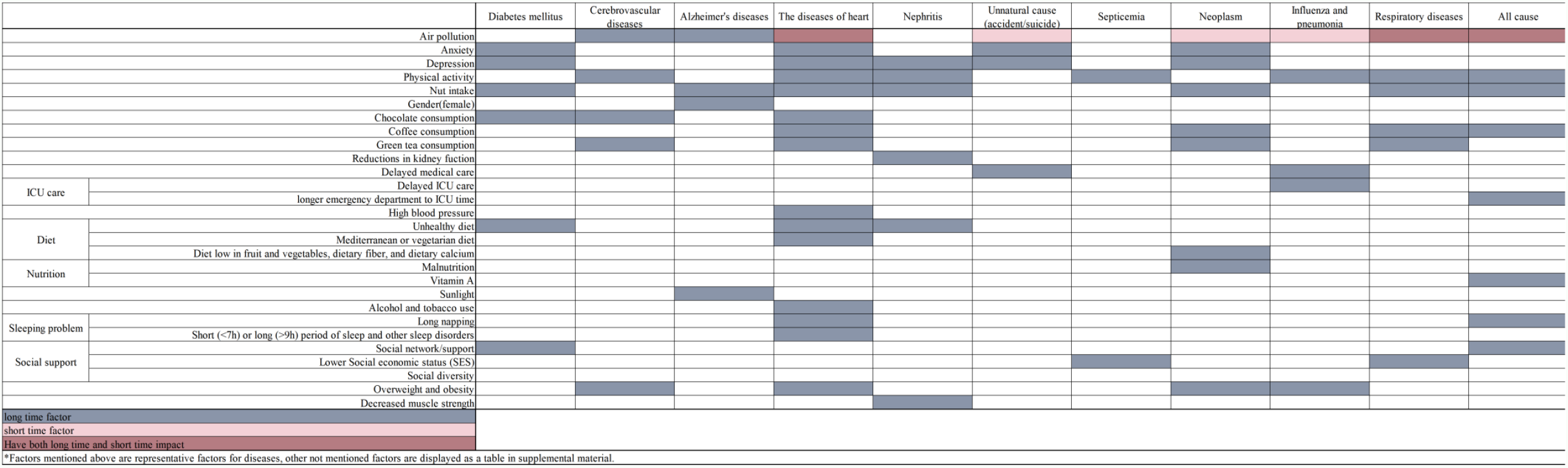
Long-term or short-term factors associated with cause-specific mortality.

### 1. Diabetes mellitus

A research investigated that the unhealthy diet is one of the drivers of the elevated morbidity of type 2 Diabetes mellitus (T2DM) which lead to the increased mortality(12). Another research in Norway indicated that compared to the reference group(no anxiety/depression symptoms), patients with anxiety symptoms had lower mortality risk (HR 0.66, 95% CI 0.46–0.96) (p = 0.03) when patients with depressive symptoms had higher mortality risk (HR 1.59, 95% CI 1.21–2.09) (p = 0.001)(13). After applying the time lag of one year, there was no positive association between moderate depressive symptoms (CES-D 16-21) and mortality risk, while there was no positive association between severe depression remains and mortality (HR=1.46, 95%CI 1.01-2.13)(14). People with medium levels of social support and the highest levels of social support have 41% (HR=0.59, 95%CI 0.39-0.91) and 55% (HR=0.45, 95%CI 0.21-0.98) lower risk of death compared with people with a low level of support(15). The meta-analysis indicated that the group with low level of support have a higher risk of mortality (HR=2.6, 95%CI 1.3-5.5)(16). Another meta-analysis revealed that the increase of nut intake decreased the risk of death caused by diabetes (RRs=0.61, 95%CI 0.43-0.88, I^2^=0%)(17). The chocolate consumption had a significant negative effect on the risk of diabetes in a research with 146,385 participants (RRs= 0.842, 95%CI 0.56-1.26, P value=0.02, I^2^=54.2%)(18). There was no short-term impact factors for this cause-specific mortality.

### 2. Cerebrovascular diseases

The study about CVD indicated that overweight (HR = 1.473, 95%CI 1.013–2.142) or obesity (HR = 1.711, 95%CI 1.1754–2.490) was associated with higher CVD mortality risk(3). Another research indicated that six air pollutants were positively associated with mortality of CVD, including PM_10_ (HR=1.11, 95%CI 1.08-1.15), PM_2.5_ (every increased 10µg/ increases 1.02 death rate), Ozone (HR=0.93, 95%CI 0.90-0.96), Nitrogen Oxides (every increased 10µg/ increases 4.5% death rate), Sulphur Dioxide (SO2)(19). The green tea consumption has a negative effect on CVD mortality in male (HR=0.82(0.75-0.90)) and female (HR=0.78(0.68-0.89))(20). Lack of exercise is a major cause of chronic diseases including CVD(21). Chocolate consumption is also a protective factor of CVD (RR=0.650, P value=0.01, 95%CI 0.15-2.74)(18).There was no short-term impact factors for this cause-specific mortality.

### 3. Alzheimer’s diseases

The female has higher risk on suffering from Alzheimer’s diseases (AD) (HR=1.7, 95%CI 1.3-2.2)(22). A research provided evidence that PM exposure can reduced cognitive abilities across the age spectrum while older ages are more obvious and higher risk of AD(23). A meta-analysis indicated that neurodegenerative disease is negatively affected by nut intake (RRs=0.65, 95%CI 0.40-1.08, =5.9%)(17). Low average sunlight (vitamin D deficiency) is also a risk factor of mortality from AD(24). There was no short-term impact factors for this cause-specific mortality.

### 4. Heart diseases

The study from US CDC has concluded many potential factors influencing the risk of heart diseases and some are: high blood pressure, obesity, diet (eating a diet high in saturated fats, trans fats, and cholesterol), alcohol and tobacco use(7). All factors mentioned above are risk factors. Air pollution also positively affects mortality from heart diseases. PM_10_ increases the risk of cardiopathy by 1.06 (95%CI 1.04-1.08) while PM_2.5_ increases even more. However, the Ozone can reduce themortality risk from heart diseases. Every increasing 10µg/ nitrogen oxides causes the risk of hospitalization rate increases 3.5%. Sulphur dioxide (SO_2_) and carbon monoxide (CO) can also positively affect the mortality of heart diseases(19). A research revealed that HADS depression sub scale≥ 8 (age, congestive heart failure, left ventricular ejection fraction, 3-vessel disease, renal disease adjusted) were significantly associated with increased risk of heart diseases (HR=2187, 95%CI 1.47-3.22, P-value < 0.001). Patients suffer from anxiety (HR=1.83, 95%CI 1.18-2.83, P=0.006) or depression (HR=1.66, 95%CI 1.06-2.58, P=0.025) or both two factors (HR=3.10, 95%CI 1.95-4.94, P<0.001) also had higher mortality(25). Short (<7h) or long (>9h) period of sleep and other sleep disorders also acted as risk factor(26). The long-time napping causes the increasing of heart diseases risk [45]. The moderate consumption of coffee (2-4 cups/day) is able to reduce the mortality risk of heart disease (27). The green tea is also a good choice for protecting drinker from heart disease for both man (HR=0.82(0.75-0.90)) and woman (HR=0.75(0.68-0.84))(20). A meta-analysis indicated that every increase 28g/d nut intake reduces the mortality of CVD to 0.71 (95%CI 0.61-0.80, *I*2=47%)(17). A research including 240 articles revealed that chocolate consumption reduce the risk of CVD and acute myocardial infarction(18). Multiple articles reveal that moderate physical activity (adjusted RR=0.71, 95%CI 0.51-0.99) and high physical activity (adjusted RR= 0.56, 95%CI 0.38-0.82) act as protective factors for coronary heart diseases(28). Meanwhile, engaging in guidelines-recommended muscle strengthening activity (HR=0.89, 95%CI 0.85-0.94) or aerobic activity (HR=0.71, 95%CI 0.69-0.72) reduce mortality of cardiovascular diseases (n=13509)(8). Diet can also play a protective factor for heart diseases. Mediterranean diet had been confirmed negative effect on CVD-related mortality(29). Vegetarian dietary pattern can reduce the risk of CHD by 40% while plant-based diets showed reversal of it (30).

Air pollution is a short-term impact factor as well. Evidence showed that each increase of 10g/ PM_2.5_ (estimate risks ER=68%, 95%CI 0.39-0.97%), PM_10_ (ER=0.39%, 95%CI 0.26-0.53%), NO_2_ (ER=1.12%, 95%CI 0.76-1.48%), SO_2_ (ER=0.75%, 95%CI 0.42-1.09%), O_3_ (ER=0.62%, 95%CI 0.33-0.92) positively affects the incidence of mortality from CVD. It also lags 0, 1, 2, 0-1, 0-2 days of exposure among pollutants and the strongest association occurred at lag 0-1day(31).

### 4. Nephritis, nephrotic syndrome, and nephrosis

The risk factors on Nephritis, nephrotic syndrome, and nephrosis include physical activity, nutrition, reductions in kidney function, depression and nut intake. Reductions in kidney function is associated with higher risk of mortality(4, 32). A review including 16 studies proved that the absolute risk of death increased exponentially as kidney function declined(32). By using the Beck Depression Inventory (BDI) and the National Institute of Mental Health Diagnostic Interview Schedule (DIS) as indicators, the point prevalence of depression is respectively 54.8% and 21.6% in severe chronic kidney disease (CKD), while 32.8% and 13.0% in the absence of severe CKD (33). Low-protein diet may probably affect the long-term renal protection. The risk of End-stage kidney disease (ESKD) increases linearly with increased protein intake(34). Decreased muscle strength is associated with increased kidney disease mortality (HR=1.84, 95% CI 1.37-2.47). On the contrary, 5 kg increased in muscle strength shows association with a decrease in kidney disease (HR=0.8, 95%CI 0.73-0.91)(35). There is an inconspicuous inverse association between increasing nut intake by one serving per day (one serving = 28 grams) and kidney disease mortality(17). There was no short-term impact factors for this cause-specific mortality.

### 6. Unnatural cause(Accident/suicide)

The risk factors includes anxiety and depression. Compared with the general population, people with anxiety disorders have a higher risk of unnatural death (Unnatural MRR=2.46, 95%CI 2.20-2.73)(36). Delays in transfers of patients to the hospital and lacking of pre-hospital emergency services is significantly associated with mortality (P<0.001)(37). In the general population, the air pollution is positively associated with increased risk of suicide death(38). The short-term risk factors associated with unnatural cause are medical care and air pollution.

### 7. Septicemia

The long-term risk and protective factors for septicemia are social economic status and physical activity. The counties with strongly clustered sepsis are characterized as higher unemployment rate, larger population in poverty, lower household income and lack of insurance coverage(39). Lack of physical activity has a negative effect on mortality of septicemia. A study shows that inadequate physical activity is associated with a 2.13-fold greater risk for sepsis mortality(40). There was no short-term impact factors for this cause-specific mortality.

### 8. Neoplasm

The long-term risk factors for neoplasm are nutrition, alcohol consumption, anxiety/depression, obesity, physical activity, coffee/green tea consumption, socioeconomic status, nut intake and medical care. Eating red and processed meat, diet with low fruits and vegetables, dietary fiber, and dietary calcium may positively affect neoplasm deaths(5). Meanwhile, epidemiological data indicates that coffee/green tea consumption(20, 41), nut and whole-grain intake(17, 42)are negatively related to the risk of neoplasm. Moreover, moderate fish consumption has negative correlation with liver cancer(43). Malnutrition has a negative effect on neoplasm survival rate. Lacking of proper nutritional management may limit the response to the most effective therapy(44).Depression and anxiety are associated with significantly increased risk of cancer-specific mortality (RR=1.21, 95%CI: 1.16–1.26) (45). The obesity and obesogenic diets accelerate the development of cancer and metastasize at an earlier age(6).

The short-term risk factor is air pollution. Air pollution, including PM_2.5_, PM_10_, O_3_, CO, SO_2_, and NO_2_/NO_x_, is confirmed as a risk factor of neoplasm mortality with time lag of less than one month (19).

### 9. Influenza and pneumonia

A meta-analysis showed that patients with overweight presented higher mortality of influenza (OR=1.99, 95%CI 1.15-3.46, I^2^=82.7%, n=7). Moreover, excessive obesity significantly increased the death of influenza (OR=1.40, 95%CI 1.10-1.79, I^2^=81.7%, n=7) (46). Delayed medical attention (OR=13.91, 95%CI 1.09–41.42, p = 0.044) and ICU hospitalization (OR=11.02, 95%CI 1.59–76.25, p=0.015) can affect influenza A/H1N1 mortality(47). People who both engaged in muscle strengthening activity and aerobic activity can present 54% lower risk of influenza and pneumonia (HR=0.46, 95%CI 0.32-0.64) compared with people not participated in physical activities(8).

With regards to short-term factors with time lag of shorter than one-month, exposure to PM_2.5_ concentrations above 12 μg/ within 4-5 days and NO_2_ within 5-6 days have a significant association with pneumonia mortality in a study with time lag adjustment(48).

### 10. Other Respiratory diseases

A systematic review about the effect of air pollution on mortality revealed that there are six air pollutants (PM_2.5_, PM_10_, O_3_, CO, SO_2_, and NO_2_/NO_x_) increasing respiratory death(19). In a cohort study in England, Ozone can influence respiratory disease mortality (HR=0.93, 95%CI 0.90-0.96) as a protective factor(49). Besides, exposure to SO_2_ and CO air-pollution can increase respiratory mortality(50). As for food consumption and intake, a meta-analysis showed a increase of 28 grams/day in nut intake reduces deaths of other respiratory disease (RR=0.48, 95%CI 0.26–0.89, I^2^ = 61%)(17). Coffee consumption can be a protective factor of mortality from other respiratory disease in another study(51). 3-4 cup green tea per day (HR=0.75 (0.61– 0.94)) and ≥ 5 cups per day (HR=0.66(0.55–0.79)) can decrease mortality from the other respiratory disease(20). A cohort study showed the risk of chronic lower respiratory tract diseases mortality was found to be 71% (HR=0.29, 95%CI 0.23-0.37) lower in those who both participated in muscle strengthening activity and aerobic activity compared with participants without physical activities(8). Lower social economic status (SES) can also cause negative influence on mortality from the other respiratory diseases(52).

A research showed short term PM_2.5_ have a 2.5% (95% CI, 1.5%-3.5%) increase in chronic obstructive pulmonary disease (COPD) death(53). Five air pollutants included PM_10_, PM_2.5_, SO_2_, NO_2_, and O_3_ with an increase of 10 μg/m3 moving average concentrations in the 8-day which separately corresponded to 0.18 (95% CI: 0.10, 0.26), 0.21 (95% CI: 0.11, 0.31), 0.16 (95% CI: 0.04, 0.30), 0.43 (95% CI: 0.07, 0.90), and 0.10 (95% CI: −0.08, 0.31) raise in the daily death from the other respiratory diseases. The influence of air pollution lasted 9 days (lag 0–8) on acute lower respiratory tract infections in Shenyang(54).

### 11. All-cause mortality

The air pollution was associated with the increased all-cause mortality (19). The research in France showed per 10 μg/ increase in PM_2.5_ pollutant can cause the increased risk of 1.02 for all-cause mortality(55). In a cohort study in England, Ozone affected all-cause death (HR=0.96, 95%CI 0.93-0.98) as a protective factor(49), but in some studies Ozone acted as a risk factor increasing 2% all-cause mortality(56). A study in Italy analyzed that all-cause mortality can increase 0.12% per 1 mg/ increase in CO (57). Additionally, exposure to SO_2_ and nitrogen oxides pollution were discovered to increase the risk of all-cause death. A longer time from Emergency to ICU (> 2.4 hour) is associated with increased the hospital mortality among the most severe patients(58). A study revealed taking over-60-minutes-per-day naps significantly led to a higher risk of all-cause mortality than those who took a short nap (long nap: HR=1.30, 95%CI 1.12–1.47; short nap: HR=1.09, 95%CI 1.01–1.16)(27). According to social support such as interpersonal relationships, social diversity, friend, neighbor and family-focused networks and community, more restricted networks are associated to higher mortality(59). Besides, there is a significant relationship between network type and mortality mainly in the elderly (> 70 years old). Those who live in diverse networks, friend-focused networks and community–clan networks in more limited degree, indicated a lower risk of all-cause death 7 years later(60). A meta-analysis on coffee consumption demonstrated the greatest reduction on all-cause death at consumption of 6.5 cups/day in young people (<60 years old) (RR=0.78, 95%CI 0.75–0.82) and 4.5 cups/day in the elderly (≥60 years old)(RR=0.85, 95%CI 0.80–0.91), compared with no coffee consumption(41). Another meta-analysis reported nut intake of per 28 grams/day can decrease all-cause mortality (RR=0.78, 95%CI 0.72–0.84, I^2^ = 66%, P-heterogeneity < 0.0001)(17). Additionally, vitamin A supplementation had a higher reduction on all-caused death (RR=0.88, 95%CI 0.83-0.93)(61). The research on physical activities presented 11% lower risk of all-cause mortality in those participating in muscle strengthening activity (HR=0.89, 95%CI 0.85-0.94) and 29% lower in those engaging in aerobic activity (HR=0.71, 95%CI 0.69-0.72) and 40% lower in those who attending both activities (HR=0.60, 95%CI 0.57-0.62)(8).

In aspect of short-term factor, a meta-analysis showed 0.89% increase in all-cause mortality per 10 µg/ increase in PM_2.5_ in previous one day (95%CI 0.68-1.10%, number of cities=114)(62).

## Discussion

We searched the PubMed dataset for researches published in English language and summarized as many as possible literature in the past 15 years of the pre-COVID-19 period which avoid the impact of pandemic. We summarized the potential factors associated with the all-cause mortality and ten main cause-specific mortality. We divided the potential factors with positive or negative effect on mortality, we also classified the potential factors with long-term (no evidence for time lag of less than on month) or short-term effect (with evidence for time lag of less than on month) on all-cause mortality and ten main cause-specific mortality.

### 1. Overall effect of potential factors associated multiple mortalities

There are several factors associated with multiple cause-specific mortality. The nut consumption could decrease the mortality risk from heart diseases, cardiovascular disease, cancer, diabetes, Nephritis, nephrotic syndrome, and nephrosis, other respiratory diseases and all-cause mortality. The moderate nuts intake is capable to protect people multi-organ, multi-system, multiple type or multiple incentives diseases.Therefore, diet suggestions on increasing nuts intake should be improved to decrease the mortality risk for human beings (17). Similarly, the factors including the physical activity, chocolate/coffee/green tea consumption, social network/support, have significant association with mortality risks from multiple causes. There were some evidences that if the factors have been changed, the mortality risk could decrease(7). The study in Mexico indicated the connection between change of influenza A/H1N1 and improved factors: encourage people to engage in physical exercise and urge people to find medical assistance initially when people have infection of influenza(47). The psychological factors may have different effect on various mortalities. The anxiety was associated with the decrease of the mortality of diabetes mellitus. Interestingly, the depression was associated with the increase of mortality of diabetes mellitus. Majority of air pollutant (PM_2.5/10_, SO_2_, NO_x_) are risk factors for cerebrovascular diseases while ozone is the protective factor either in short term or long term. In other diseases, however, all air pollutants show the same positive influence.

### 2. Protective and risk factors associated with cause-specific mortality

Our review showed that there are a variety of factors can affect the cause-specific diseases significantly. For those risk factors, more researches were further required to investigate the effect of decreasing such negative factors on mortality risk.

For instance, obesity, as a risk factor, significantly increased the mortality risk of cerebrovascular disease, heart diseases, neoplasm, influenza and pneumonia, therefore, weight loss should be encouraged by the governments as an important public health interventions for mortality control. At the same time, more studies about the effect of controlling body weight on mortality risk of specific diseases are of great importance. Besides, more researches about effective methods for weight loss should be considered in the future. For protective factors, such as nuts intakes, coffee or green tea consumption, physical activities, more high-quality researches should be implemented to investigate the impact of increase on above protective factors for human beings on mortality risk.

### 3. Long-term or short-term factors associated with cause-specific mortality

With regard to analyzing the long-term or short-term impact of factors, only air pollution have short-term effect on the mortality of heart disease, unnatural cause(accident/suicide), neoplasm, influenza and pneumonia, respiratory disease, all cause. Short-term PM_2.5_ increase the specific diseases mortality as a rapid factor, therefore, it is a warning for patients to avoid contacting polluted air to raise the quantity of live and be longevity. People are suggested to realize the threat of long-term impact factors should be focused on to engaged in a long-term beneficial behavior, such as healthy diet, physical activities.

## 4. Limitations

Articles for our review is collected during 2004 to 2021 mainly from PubMed website and only English articles were included in this review, therefore, our study may miss some potential factors published in other languages. considered factors were either notable significantly affecting the mortality or revealing a potential connection with the mortality therefore some factors possibly not consider in the review. Source data of mortality in reference were not identical which is beyond our control. Consequently, the mortality of diseases has slight distinctness from article to article. Identically, owing to the differences of scope, influence caused by nations, region, race is not considered and adjusted.

## 5. Conclusion

Generally, our review displayed various factors have a positive or negative and long-term or short-term effect on cause-specific disease mortality. As the review is to research pre-COVID-19 period time, whether COVID-19 is a significantly direct factor for the infection and death of these diseases or not require further researches. After acquiring a variety of significant factors influenced on specific disease death, people can engage in protective factors and avoid risk factors to lower the probability of illness, raise the quality of life and decrease the mortality of multiple disease.

## Supporting information

supplementary material-review

## Data Availability

All data produced in the present work are contained in the manuscript.

