## supplementary material-review for "The effect of potential factors on all-cause and cause-specific and mortality: a pre-COVID-19 period review"

### Supplemental Appendix

**Table S1. Summary of potential drivers associated with mortality described in the pre-COVID-19 pandemic literature**

| Deaths | Factors related to deaths | Association | Lag | Data |
|---|---|---|---|---|
| **Diabetes mellitus** | Unhealthy diet[1] | Positive | | |
| | Depression[2] | Positive | 1 year[3] | √ |
| | Smoking and drinking alcohol[4] | Positive | | |
| | Complication (cardiovascular complications)[2,4,5] | Positive | | |
| | Soya and palm oils[6] | Positive | | |
| | Social support*[7,8] | Negative | | |
| | Socioeconomic status[9] | Negative | | |
| | Nut intake[10] | Negative | | |
| | Anxiety[2] | Negative | | √ |
| | Sedentary lifestyle/physically active[3,11] | Negative | | |
| | Chocolate consumption[12] | Negative | | |
| | Quality of diabetes Care[13] | Negative | | |
| | Whole-grain Intake[14] | Negative | | |
| | Sunlight[15] | Negative | | |
| **Cerebrovascular diseases** | Depressive symptoms[16] | Positive | | √ |
| | Anxiety[17] | Positive | | √ |
| | Overweight/obesity[18] | Positive | | |
| | Air pollution[19] | Positive | up to 7 days[20] | √ |
| | Obstructive sleep apnea[21] | Positive | | |
| | Soya and palm oils[6] | Positive | | |
| | Cognition function[18] | Negative | | |
| | Living in urban area[18] | Negative | | |
| | Emergency calls[22] | Negative | | |
| | Nutritional condition[18] | Negative | | |
| | Nut intake/green tea consumption[10,23] | Negative | | |
| | Physical activity[24] | Negative | | |
| | Neighborhood cohesion[25] | Negative | | |
| | Chocolate consumption[12] | Negative | | |
| | Telemedicine(through systolic blood pressure)[26] | Negative | | |
| | Fish consumption[27] | Negative | | |
| | Green space[28] | Negative | | |
| | Apple consumption[29] | Negative | | |
| **Alzheimer's diseases** | Gender[30] (female) | Positive | | |
| | Air pollution[31] | Positive | | √ |
| | Low average sunlight (vitamin D deficiency)[32] | Positive | | |
| | Presence of pressure ulcers[33] | Positive | | |
| | Comorbidities (diabetes, CVD)[33] | Positive | | |
| | Longer duration of illness[34] | Negative | | |
| | Observed depression[33] | Negative | | √ |
| | Presence of physical illness[34] | Negative | | |
| | Poor cognitive function[34] | Negative | | |
| | Nut intake[12] | Negative | | |

**Supplement Table 2 (continued). Summary of potential drivers associated with mortality described in the pre-COVID-19 pandemic literature**

| Deaths | Factors related to deaths | Association | Lag | Data |
|---|---|---|---|---|
| **The diseases of heart** | High blood pressure[35] | Positive | | |
| | Heat and cold[36] | Positive | | |
| | Diabetes[35] | Positive | | |
| | Obesity[35] | Positive | | |
| | Diet (eating a diet high in saturated fats, trans fats, and cholesterol)[35] | Positive | | |
| | Alcohol and tobacco use[35] | Positive | | |
| | Heat and cold [37] | Positive | | |
| | Air pollution[19] | Positive | 0-2 days[38] | √ |
| | Genetic factors (family history of heart disease)[35] | Positive | | |
| | Anxiety/depression[39] | Positive | | √ |
| | Loneliness[40] | Positive | | |
| | Short (<7h) or long (>9h) period of sleep and other sleep disorders[41] | Positive | | |
| | Napping[42] | Positive | | |
| | Social network/support*[43] | Negative | | |
| | Coffee consumption/green tea consumption[44,23] | Negative | | |
| | Telemonitoring and telephone interventions[45] | Negative | | |
| | Socioeconomic status[8] | Negative | | |
| | Nut intake[10] | Negative | | |
| | Hospital volume[46] | Negative | | √ |
| | Physical activity[35,47,48] | Negative | | |
| | Sauna bathing[49] | Negative | | |
| | Chocolate consumption[12] | Negative | | |
| | Mediterranean diet[50] | Negative | | |
| | Vegetarian dietary[51] | Negative | | |
| | Dairy products[52] | Negative | | |
| | Sunlight[15] | Negative | | |
| | Whole-grain Intake[14] | Negative | | |
| | Fish consumption[27] | Negative | | |
| | Olive oil Intake[50] | Negative | | |
| | Apple[26] | Negative | | |
| **Nephritis** | Elevated rates of hospitalization associated with reductions in kidney function, and participants with reduced kidney function have higher risk of mortality [54,55] | Positive | | |
| | Poor mental health [56] | Positive | | √ |
| | Depression[57] | Positive | | √ |
| | Physical activity [58] | Negative | | |
| | Nut intake[10] | Negative | | |
| | Muscular strength[59] | Negative | | |
| | Protein restriction (for end-stage kidney disease) [60] | Negative | | |

**Supplement Table 2 (continued). Summary of potential drivers associated with mortality described in the pre-COVID-19 pandemic literature**

| Deaths | Factors related to deaths | Association | Lag | Data |
|---|---|---|---|---|
| **Unnatural cause<br><br>-accident<br>-suicide** | Air pollution ($NO_2$, $SO_2$, $PM_{10}$, and $PM_{2.5}$)[61] | Positive (suicide) | 0-1 day[58] | √ |
| | Anxiety[62] | Positive | | √ |
| | Depression[62] | Positive | | √ |
| | Choose home for suicide[63] | Positive | | |
| | Airborne pollen (for women in suicide)[61] | positive | 0 day[64] | |
| | Traffic noise[65] | Positive (suicide) | 1 day[65] | |
| | Higher rate of bank suspensions[66] | Positive (suicide) | | |
| | Higher rate of bank suspensions[66] | Negative (traffic accident) | | |
| | Delays in transfers of patients to the hospital, and a lack of pre-hospital emergency services[67] | Negative | | |
| **Septicemia** | Rural population[68] | Positive | | |
| | Unemployment rate[68] | Positive | | |
| | Comorbidities[69,70] | Positive | | |
| | Organ failure[70] | Positive | | |
| | Race[68](black) | Positive | | |
| | Poverty[68] | Positive | | |
| | Disability in activities[71] | Positive | | |
| | ICU care[69] | Negative | | √ |
| | Household income | Negative | | |
| | Physical activities[72] | Negative | | |
| | Insurance coverage[69] | Negative | | |
| **Neoplasm** | Depression[73] | Positive | 1 year[74] | √ |
| | Obesity[75] | Positive | | |
| | Anxiety[73] | Positive | | √ |
| | Air pollution[19] | Positive | (Lung cancer)<br>3 weeks[76] / 0-8 year[77,78];<br>(Stomach/colorectal<br>Cancer)<br>0-7 days[79] | √ |
| | Drinking alcohol[80] | Positive | | |
| | Eating red and processed meat[80] | Positive | | |
| | Diet low in fruits and vegetables, dietary fiber, and dietary calcium[80] | Positive | | |
| | Six cancer-associated infections – Helicobacter pylori, hepatitis B virus (HBV), hepatitis C virus (HPC), human herpes virus type 8 (HHV8), human immunodeficiency virus (HIV), and human papillomavirus (HPV) [80] | Positive | | |
| | Ultraviolet (UV) radiation from the sun or indoor tanning[80] | Positive | | |
| | Malnutrition[81] | Positive | | |
| | Red and processed meat[82] | Positive | | |
| | Physical activity[48] | Negative | | |
| | Coffee/green tea consumption[23,44] | Negative | | |

**Supplement Table 2 (continued). Summary of potential drivers associated with mortality described in the pre-COVID-19 pandemic literature**

| Deaths | Factors related to deaths | Association | Lag | Data |
|---|---|---|---|---|
| **Neoplasm** | Socioeconomic status[9] | Negative | | |
| | Nut intake[10] | Negative | | |
| | Muscular strength[59] | Negative | | |
| | Medical care (screening, diagnosis, and treatment)[83] | Negative | | |
| | Whole-grain Intake[14] | Negative | | |
| | Fish consumption (liver cancer)[27] | Negative | | |
| **Influenza and pneumonia** | Obesity[84] | Positive | | |
| | Medical attention delayed[85] | Positive | | |
| | Air pollution[86, 87] | Positive | (Pneumonia) 0-6 days[86];(Acute lower respiratory tract infection) 0-8 days[87] | √ |
| | Admission to ICU[85] | Positive | | √ |
| | Creatinine levels when admitted to hospital[85] | Positive | | |
| | Pandemic influenza hospitalization was more common in individuals with type 2 diabetes[88] | Negative | | √ |
| | Physical activity[85,48] | Negative | | |
| | High-dose vitamin C therapy[88] | Negative | | |
| **Respiratory diseases** | Anxiety | Positive | | √ |
| | Driving pressure[90] | Positive | | |
| | Heat and cold[37] | Positive | | |
| | Air pollution/PAH in the air[19,91] | Positive | (COPD) 0-7 days[92] | √ |
| | Social Economic Status (medical care) | Negative | | |
| | Treatment with noninvasive oxygenation strategies | Negative | | |
| | Non-invasive ventilation | Negative | | |
| | Nut intake[10] | Negative | | |
| | Coffee consumption/green tea consumption[93,23] | Negative | | |
| | Muscular Strength[59] | Negative | | |
| | Physical activity[48] | Negative | | |
| **All cause** | Air pollution[19] | Positive | 1 day[94] | √ |
| | Emergency department to ICU Time[95] | Positive | | |
| | Napping[42] | Positive | | |
| | Social network (interpersonal relationships)[43] | Negative | | |
| | Coffee consumption[44] | Negative | | |
| | Nut intake[10] | Negative | | |
| | Vitamin A[96] | Negative | | |
| | Social diversity, friend-focused, neighbor-focused, family-focused, community[97] | Negative | | |
| | Fish consumption[27] | Negative | | |
| | Whole-grain intake[14] | Negative | | |
| | Green space[28] | Negative | | |
| | Sunlight[15] | Negative | | |
| | Apple[29] | Negative | | |

* **Social support:** quantitative or social network support, referring to the number of persons to whom someone can resort for assistance or in case of need; and qualitative or functional support of a subjective nature, referring to each individual's perception of availability of support.
